# Subacute effects of the psychedelic ayahuasca on the salience and default mode networks

**DOI:** 10.1101/19007542

**Authors:** Lorenzo Pasquini, Fernanda Palhano-Fontes, Draulio B. Araujo

**Author notes:** These authors contributed equally to the manuscript (Lorenzo Pasquini, Fernanda Palhano-Fontes). Corresponding author: Draulio B. Araujo, Brain Institute (UFRN), Av. Nascimento Castro, 2155, Natal, RN 59056 - Brazi.

## Abstract

**Background:** Neuroimaging studies have just begun to explore the acute effects of psychedelics on large-scale brain networks’ functional organization. Even less is known on the neural correlates of subacute effects taking place days after the psychedelic experience. This study explores the subacute changes of primary sensory brain networks and networks supporting higher-order affective and self-referential functions 24h after a single session with the psychedelic ayahuasca.

**Methods:** We leveraged task-free functional magnetic resonance imaging data one day before and one day after a randomized placebo-controlled trial exploring the effects of ayahuasca in naïve healthy participants (21 placebo/22 ayahuasca). We derived intra- and inter-network functional connectivity of the salience, default mode, visual, and sensorimotor networks, and assessed post-session connectivity changes between the ayahuasca and placebo groups. Connectivity changes were associated with Hallucinogen Rating Scale scores assessed during the acute effects.

**Results:** Our findings revealed increased anterior cingulate cortex connectivity within the salience network, decreased posterior cingulate cortex connectivity within the default mode network, and increased connectivity between the salience and default mode networks one day after the session in the ayahuasca group compared to placebo. Connectivity of primary sensory networks did not differ between-groups. Salience network connectivity increases correlated with altered somesthesia scores, decreased default mode network connectivity correlated with altered volition scores, and increased salience-default mode network connectivity correlated with altered affect scores.

**Conclusion:** These findings provide preliminary evidence for subacute functional changes induced by the psychedelic ayahuasca on higher-order cognitive brain networks that support interoceptive, affective, and self-referential functions.

## Introduction

Classic psychedelics belong to a family of substances that lead to altered states of consciousness, with a broad range of effects on sensory, cognitive, autonomic, interoceptive, self-referential, and emotional systems (Nichols, 2016). Assisted sessions with psychedelic substances such as ayahuasca and psilocybin have recently gained prominent use for the treatment of affective disorders (Carhart-Harris, Bolstridge, et al., 2016; Griffiths et al., 2016; Grob et al., 2011; Osório et al., 2015; Palhano-Fontes et al., 2019; Ross et al., 2016; Sanches et al., 2016).

Ayahuasca is an indigenous preparation that contains leaves of *Psychotria viridis* and the bark of *Banisteriopsis caapi*, two plants vastly found in the Amazon basin (Mckenna et al., 1984). The admixture contains the psychedelic tryptamine N,N-dimethyltryptamine (N,N-DMT) and monoamine oxidase inhibitors. The first has a particularly high affinity to serotonin and sigma-1 receptors while the latter renders N,N-DMT orally active by inhibiting the enzyme monoamine oxidase in the gut (Barker, 2018; Fontanilla et al., 2009; Mckenna et al., 1984; Riba, Rodríguez-Fornells, et al., 2001). Recent clinical trials have explored the antidepressant effects of ayahuasca (Osório et al., 2015; Palhano-Fontes et al., 2019; Sanches et al., 2016). In an open-label trial, 17 patients with treatment-resistant depression were submitted to a single session with ayahuasca. Significant antidepressant effects were observed one day after the session and persisted for 21 days (Osório et al., 2015; Sanches et al., 2016). Consistent results were found in a separate randomized placebo-controlled trial, where a significant rapid antidepressant effect of ayahuasca was observed already one day after the session, compared to placebo (Palhano-Fontes et al., 2019). Yet, little is known about the underlying neural mechanisms by which ayahuasca may modulate affect, mood, and internal representations of oneself.

Neuroimaging techniques provide a powerful tool to explore brain changes elicited by psychedelic agents (Dos Santos et al., 2016; Dos Santos et al., 2020). Task-free functional magnetic resonance imaging (tf-fMRI, also referred to as resting-state fMRI) measures synchronous, low frequency (<0.1 Hz) fluctuations in blood oxygen level-dependent (BOLD) activity among distant gray matter regions (Allen et al., 2011; Fox et al., 2005; Smith et al., 2009) and has been used to delineate distinct intrinsic large-scale brain networks supporting primary sensory and higher cognitive and emotional functions (Fox et al., 2005; Grayson and Fair, 2017; Smith et al., 2009; van den Heuvel and Hulshoff Pol, 2010). Importantly, human emotions and social behavior are modulated by a distributed set of brain regions including the anterior cingulate cortex, bilateral anterior insula, and subcortical and limbic structures that regulate affect, interoception, and self-awareness by interacting with the autonomic nervous system (Craig, 2009; Critchley and Harrison, 2013; Lorenzo Pasquini et al., 2019; L. Pasquini et al., 2019; Zhou and Seeley, 2014). These regions coactivate and are structurally connected, cohesively forming the salience network, a large-scale brain system involved in homeostatic behavioral guidance and supporting socioemotional functions by integrating visceral and sensory information (Rankin et al., 2006; Seeley et al., 2007; Sturm, Brown, et al., 2018; Uddin, 2015). A second prominent large-scale brain system is the default mode network, which is primarily composed of the posterior cingulate cortex, precuneus, inferior parietal lobe, parahippocampal gyrus, and medial prefrontal cortex (Buckner et al., 2008). This network has been repeatedly associated with self-referential processes such as mind-wandering, introspection, and memory retrieval under healthy and pathological conditions (Buckner et al., 2008; Raichle et al., 2001).

Recent neuroimaging studies have explored the acute effects of psychedelics on functional organization of human brain networks (Carhart-Harris et al., 2012; Carhart-Harris, Muthukumaraswamy, et al., 2016; Palhano-Fontes et al., 2015; Roseman et al., 2014; Viol et al., 2017, 2019), with reports of increased functional integration at the global brain level (Petri et al., 2014; Tagliazucchi et al., 2016; Viol et al., 2017), and connectivity increases primarily affecting the primary visual cortex and frontal areas (Carhart-Harris, Muthukumaraswamy, et al., 2016; De Araujo et al., 2012). Only one study has reported salience network connectivity decreases under acute conditions (Lebedev et al., 2015), while most studies report decreased connectivity in important hubs of the default mode network, such as the posterior cingulate cortex (Carhart-Harris et al., 2012; Lebedev et al., 2015; Palhano-Fontes et al., 2015).

A recent study assessing subacute neural changes 24 hours after a single dose of ayahuasca reported decreased negative connectivity between the anterior cingulate cortex, a salience network hub, and key default mode network regions such as the posterior cingulate cortex and medial temporal lobes. These connectivity changes correlated with increased self-compassion scores and other measures of enhanced psychological capacities assessed 24 hours after the session, providing a biological basis for the post-acute or “after-glow” stage of psychedelic effects (Majić et al., 2015; Sampedro et al., 2017). Likewise, subacute effects of psilocybin in patients with treatment-resistant depression one day after dosing led to increased connectivity between hubs of the salience and default mode networks such as the subgenual cingulate and posterior cingulate cortices, while connectivity decreases were observed in patients between the prefrontal and parahippocampal cortices, two important default mode network regions (Carhart-Harris et al., 2017). Little is known, however, on inter- and intra-network subacute changes induced by ayahuasca on the salience and default mode networks of naïve healthy participants hours to days after a psychedelic session, and how functional changes may relate to the altered state of consciousness elicited during the psychedelic experience.

To address this critical gap, we leveraged tf-fMRI data assessed before and 24 hours after a randomized placebo-controlled trial exploring the subacute effects of ayahuasca in naïve healthy participants. We hypothesized that 24 hours after ayahuasca administration, we would observe changes in higher-order affective and self-referential networks but not primary sensory networks. Furthermore, we predicted that changes in higher-order affective and self-referential networks would relate to the acute effects of ayahuasca. We applied a seed-based connectivity approach to map the salience, default mode, visual, and sensorimotor networks and assessed subacute inter- and intra-network connectivity changes one day after the session with ayahuasca compared to placebo.

## Methods and Materials

We recruited 50 participants from local media and the Internet. All individuals included in the study were cognitively and physically healthy and naïve to ayahuasca. The following exclusion criteria were adopted: (i) diagnosis of current chronic disease (e.g. heart disease, diabetes) based on anamnesis, physical, and laboratory examination; (ii) pregnancy; (iii) history of neurological/psychiatric diseases; (iv) substance abuse (DSM-IV); (v) restrictions to magnetic resonance imaging assessment. All procedures took place at the Onofre Lopes University Hospital (UFRN), Natal-RN, Brazil. The study was approved by the University Hospital Research Ethics Committee (# 579.479). All subjects provided written informed consent before participation. The study was registered at http://clinicaltrials.gov (NCT02914769).

Half of the participants (n = 25) received a single low dose of 1 ml/kg of ayahuasca and the other half (n = 25) received 1 ml/kg of a placebo substance. Alkaloids concentrations were quantified by mass spectroscopy analysis. A single ayahuasca batch was used throughout the study, containing on average (mean ± S.D.): 0.36 ± 0.01 mg/ml of N, N-DMT, 1.86 ± 0.11 mg/ml of harmine, 0.24 ± 0.03 mg/ml of harmaline, and 1.20 ± 0.05 mg/ml of tetrahydroharmine. The batch was prepared and provided by a branch of the Barquinha church, Ji-Paraná, Brazil. The liquid used as placebo was designed to simulate organoleptic properties of ayahuasca, such as a bitter and sour taste, and a brownish color. It contained water, yeast, citric acid, zinc sulfate, and caramel colorant. The presence of zinc sulfate also produced modest gastrointestinal distress allowing to control for the non-specific effects of ayahuasca (Palhano-Fontes et al., 2019). Analogously to two previous studies investigating the subacute effects of psychedelics (Carhart-Harris et al., 2017; Sampedro et al., 2017), baseline tf-fMRI assessments occurred one day before dosing, and a second tf-fMRI assessment occurred 24 hours after dosing (Figure 1A). Blood samples were drawn at baseline and one day after the session.

**Figure 1.**
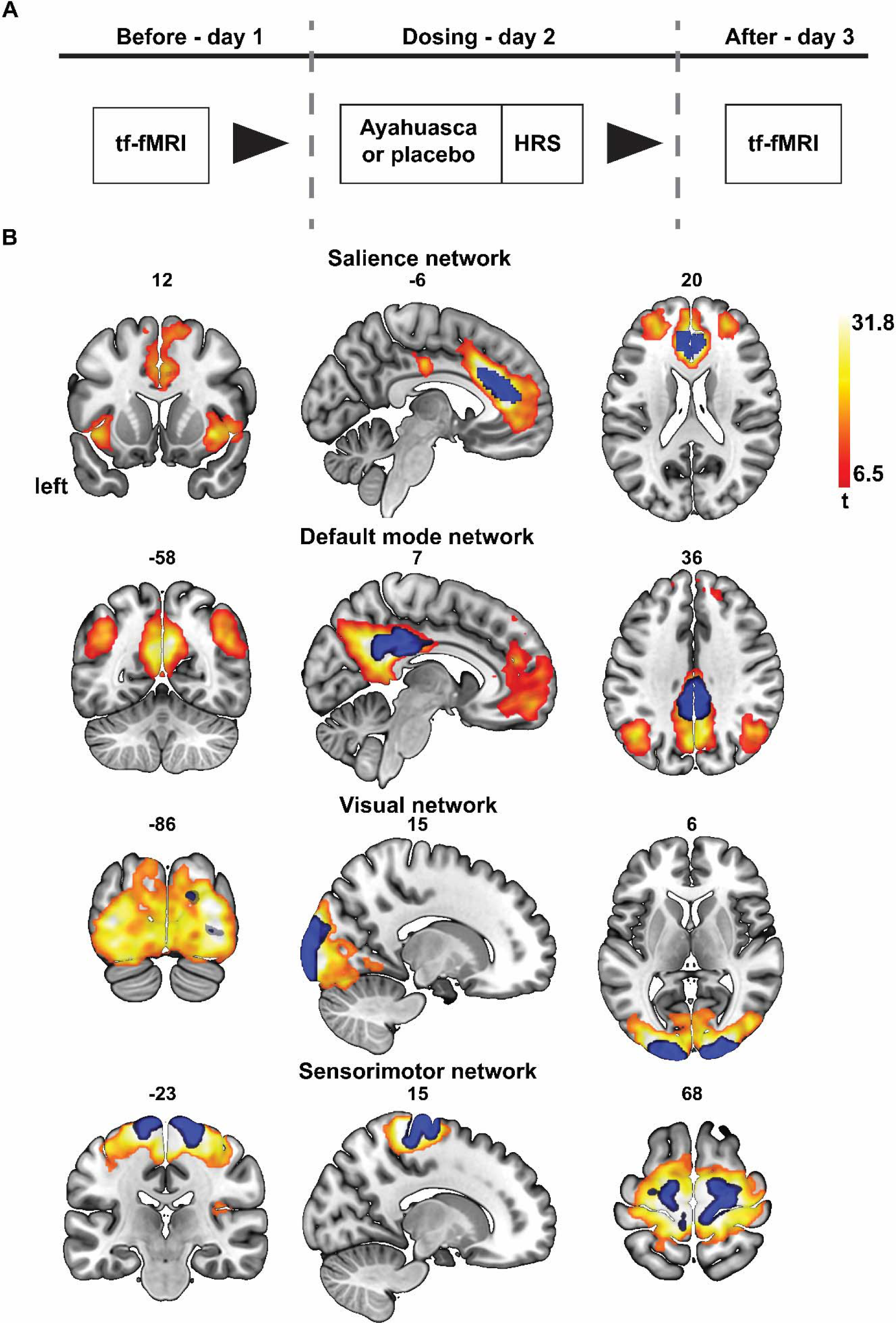
Study pipeline. **(A)** Participants were randomly assigned to the placebo (n = 21; 1 ml/kg of ayahuasca adjusted to contain 0.36 mg/kg of DMT) or ayahuasca group (n = 22; 1 ml/kg of a placebo). Subscales of the Hallucinogenic Rating Scale (HRS) were assessed during the acute effects of ayahuasca/placebo. All participants were assessed with tf-fMRI one day before and one day after dosing. **(B)** Map of the salience, default mode, visual, and sensorimotor networks at baseline overlaid onto the MNI template (warm colors; extent probability threshold of p < 0.01 FWE corrected, voxel-wise threshold of p < 0.001 FWE corrected for multiple comparisons). Individual brain network maps were derived by seeding bilateral region-of-interest (in blue). Left is on the left, bar reflects t-values.

To assess the acute psychedelic experience, we used the Hallucinogenic Rating Scale (HRS), which was initially designed to assess the psychedelic effects of N,N-DMT (Strassman et al., 1994). It is divided into six subscales: (i) intensity, which reflects the strength of the overall experience; (ii) somesthesia, which assesses somatic effects including interoception, visceral, and tactile effects; (iii) affect, which assesses comfort, emotional, and affective changes; (iv) perception, which assesses visual, auditory, gustatory, and olfactory effects; (v) cognition, which assesses changes in thoughts; and (vi) volition, which is related to the subject’s capacity to willfully interact with his/her ‘self’ (Riba, Rodri, et al., 2001; Strassman et al., 1994). We used the validated HRS version translated to Brazilian Portuguese (Mizumoto et al., 2011). Participants completed the HRS at the end of the dosing session (approximately 4 hours after ayahuasca or placebo ingestion).

### Neuroimaging data acquisition

tf-fMRI scanning was performed on a 1.5 Tesla scanner (General Electric, HDxt). Functional MRI data were acquired using an EPI sequence with the following parameters: TR = 2000 ms; TE = 35 ms; flip angle = 60°; FOV = 24 cm; matrix = 64 ⨯ 64; slice thickness = 3 mm; gap = 0.3 mm; number of slices = 35; volumes = 213. T1-weighted images were also acquired using a FSPGR BRAVO sequence with the following parameters: TR = 12.7 ms, TE = 5.3 ms, flip angle = 60°, FOV = 24 cm, matrix = 320 ⨯ 320; slice thickness = 1.0 mm; number of slices = 128.

### Neuroimaging data preprocessing

Of those 50 participants that met our initial inclusion criteria, three participants in the ayahuasca group and four in the placebo group were excluded from further analyses. One subject was excluded due to loss to follow up resulting in the missing of the second scan; one subject due to obvious scanning artifacts as missing of the parietal cortex in the field of view; one participant had to be excluded due to a cystic lesion identified after visually inspecting the anatomical scans after the session; four subjects had to be excluded due to high blood glucose levels indicative of diabetes, which were revealed to the experimentator about a week after completion of the experiment.

The first three fMRI volumes were discarded to allow T1 saturation. Preprocessing steps were performed using CONN (https://web.conn-toolbox.org/) (Whitfield-Gabrieli and Nieto-Castanon, 2012) and included: slice timing correction, head motion correction, and spatial smoothing (6-mm FWHM Gaussian kernel) (**Supplementary Figure S1**). Functional images were coregistered to the subject’s anatomical image, normalized into the Montreal Neurological Institute (MNI) template, and resampled to 2 mm_3_ voxels. Motion artifact was examined using the Artifact Detection Toolbox (ART; http://www.nitrc.org/projects/artifact_detect/). Volumes were considered outliers if the global signal deviation was superior to five standard deviations from the mean signal or if the difference in frame displacement between two consecutive volumes exceeded 0.9 mm. Physiological and other spurious sources of noise were removed using the CompCor method (Behzadi et al., 2007). CompCor is a method used for the reduction of non-physiological noise (head movement, cerebrospinal fluid) in BOLD data. Anatomical data is used to define regions of interest composed primarily of white matter and cerebrospinal fluid. Principal components are derived from these noise-related regions-of-interest and are then included as nuisance parameters within general linear models for BOLD fMRI time-series data denoising. Residual head motion parameters (3 rotation and 3 translation parameters), outlier volumes, white matter, and cerebral spinal fluid signal were regressed out. Finally, a temporal band-pass filter of 0.01 Hz to 0.1 Hz was applied. We evaluated the amount of head motion in our sample by individually estimating the head mean framewise displacement at both sessions (**Supplementary Table S1**). Participants showed head mean framewise displacement way below the widely used threshold of 0.50 mm (Carhart-Harris et al., 2017) and close to the stringent threshold of 0.25 mm recommended by experts in the field (Parkes et al., 2018; Power et al., 2012, 2014; Satterthwaite et al., 2012, 2013).

### Intra-network functional connectivity analyses

For each subject, average BOLD activity time courses were extracted using bilateral regions-of-interest from the Brainnetome Atlas (seed numbers 187 and 188 for the anterior cingulate cortex, seed numbers 175 and 176 for the posterior cingulate cortex, seed numbers 203 and 204 for the occipital cortex, and seed numbers 59 and 60 for the precentral cortex, http://atlas.brainnetome.org/) (Fan et al., 2016). Salience, default mode, visual, and sensorimotor network functional connectivity maps were generated through voxel-wise regression analyses using the extracted time-series as regressors (Seeley et al., 2007). Baseline one-sample t-test maps derived from the seed-based approach were visually explored by experts and subsequently spatially correlated with publicly available maps of the visual (R_seed_ = 0.44), sensorimotor (R_seed_ = 0.49), default mode (R_seed_ = 0.69) and salience networks (R_seed_ = 0.64) (Smith et al., 2009), and interactively compared to maps available on NeuroVault (https://neurovault.org) (Gorgolewski et al., 2015). To control for our method of choice, the seed-based approach, we replicated the analysis using group independent component analysis (ICA) implemented through the GIFT toolbox (http://icatb.sourceforge.net). Briefly, preprocessed tf-fMRI data of all subjects were decomposed into 20 spatially independent components within a group-ICA framework (Calhoun et al., 2001; Pasquini et al., 2015). Data were concatenated and reduced by two-step principal component analysis, followed by independent component estimation with the infomax-algorithm. This procedure results in a set of averaged group components, which are then back reconstructed into the subject’s space. As in the seed-based approach, baseline one-sample t-test maps derived from the ICA-based approach were spatially correlated with publicly available maps of the default mode network (R_ICA_ = 0.63) (Buckner et al., 2008) and salience network (R_ICA_ = 0.56) (Smith et al., 2009).

### Inter-network functional connectivity analyses

Average time series of resting BOLD activity were derived before and after the session from the one-sample t-test maps of the salience, visual, sensorimotor, and default mode networks identified in the seed-based analyses (**Figure 1B**). Pairwise Pearson’s correlation coefficients were used to estimate the level of functional connectivity between networks at baseline and after the ayahuasca session.

### Statistical analyses

Using individual brain network connectivity maps across both samples at baseline, a voxel-wise one-sample t-test was implemented in SPM12 (https://www.fil.ion.ucl.ac.uk/spm/software/spm12/) to assess significant functional connections to our seeds of choice (t = 6.5, extent probability threshold of p < 0.01 FWE corrected, voxel-wise threshold of p < 0.001 FWE corrected for multiple comparisons; **Figure 1B**). These maps were used as masks for subsequent voxel-wise statistical analyses. Change maps of brain network functional connectivity were derived by individually subtracting the functional connectivity maps post-dosing from the ones at baseline. Voxel-wise two-sample t-tests were used to assess differences in brain network functional connectivity between placebo and ayahuasca. Control analyses were performed by adding the difference in head mean framewise displacement between both scans as a covariate of no interest. We explicitly refrained from comparing functional connectivity maps at baseline across the two groups in accordance with the recommendation from the Consolidated Standards of Reporting Trials Group (CONSORT, http://www.consort-statement.org/), which encompasses various initiatives developed by journal editors, epidemiologist, and clinical trialists to alleviate the problems arising from inadequate reporting of randomized control trials. If not reported otherwise, all voxel-wise findings are reported at the joint cluster and extent probability thresholds of p < 0.05 and p < 0.01 uncorrected. Analogously to the intra-network connectivity approach, changes in inter-network connectivity were assessed by subtracting the baseline inter-network functional connectivity value from the post-session value. The change in inter-network functional connectivity was subsequently compared across groups via two-sample t-tests (p < 0.02 uncorrected).

Since the use of liberal statistical thresholds in neuroimaging studies has been recently shown to be prone to inflate false-positives results (Eklund et al., 2016), we additionally assessed group differences in functional connectivity through nonparametric permutation tests as implemented using the FSL package Randomise (https://fsl.fmrib.ox.ac.uk/fsl/fslwiki/Randomise) (p < 0.05 threshold-free cluster enhancement uncorrected and FWE corrected for multiple comparisons) (Smith and Nichols, 2009; Winkler et al., 2014). Two-sample t-tests were used to assess group differences in demographic variables, head mean framewise displacement, and HRS subscales (p < 0.05, corrected for multiple comparisons if not specified otherwise). Average functional connectivity changes were derived from clusters identified with the voxel-wise two-sample t-tests using an uncorrected joint cluster and extent probability threshold of p < 0.05 for the default mode network and a more stringent joint cluster and extent probability threshold of p < 0.01 for the salience network. Partial correlation coefficients corrected for the difference in head mean framewise displacement between the second and first scan were used to associate intra- and inter-network functional connectivity changes with HRS subscale scores (p<0.05 if not specified otherwise).

### Data and Code availability

The data that support the findings of this study and the code used to generate the findings are available on request from the corresponding author D.B.A. Statistical t-maps shown in this study are publicly available on NeuroVault (Gorgolewski et al., 2015) (https://identifiers.org/neurovault.image:312874)(https://identifiers.org/neurovault.image:312875). The data are not publicly available due to them containing information that could compromise the privacy of participants in the study.

## Results

Forty-three (43) ayahuasca naïve participants assigned either to ayahuasca (n = 22) or placebo (n = 21) passed our neuroimaging data quality check (**Table 1**). All participants were assessed with tf-fMRI one day before and one day after dosing (**Figure 1A**), and salience, default mode, visual, and sensorimotor networks maps were individually derived by seeding bilateral regions-of-interest (**Figure 1B, Supplementary Table S2**). Participants were comparable in terms of age, gender distribution, and head mean framewise displacement at both sessions (**Table 1**). We found significant between-group differences for all HRS subscales, with significant increases in the ayahuasca compared to the placebo group in levels of altered somesthesia (p < 0.0005), affect (p < 0.005), perception (p < 0.0005), cognition (p < 0.0005), volition (p < 0.009), and intensity (p < 0.0005) (**Table 2**). All HRS subscale comparisons survived multiple correction testing (p < 0.008) with the exception of the volition subscale.

**Table 1.** Sample characteristics. ^&^Chi-square statistics, chi-square test used to test group differences in gender distribution. Paired t-test used to test differences in head motion between the first and second sessions: ^A^t = 1.43, p = 0.17; ^B^t = 0.41, p = 0.69.

|  | **Ayahuasca**<br>**(N = 22)** | **Placebo**<br>**(N = 21)** | **t** | **p** |
| --- | --- | --- | --- | --- |
| **Age (s.d.) in years** | 30.8 (8.4) | 31.0 (10.5) | 0.06 | 0.95 |
| **Gender (F/M)** | 12/10 | 11/10 | 0.02<sup>a</sup> | 0.89 |
| **Before dosing - head mean**<br>**framewise displacement (s.d) in mm** | 0.14 (0.03) | 0.13 (0.05) | 0.56 | 0.57 |
| **After dosing - head mean framewise**<br>**displacement (s.d) in mm** | 0.15 (0.05)<sup>a</sup> | 0.14 (0.05)<sup>b</sup> | 1.01 | 0.31 |

**Table 2.**
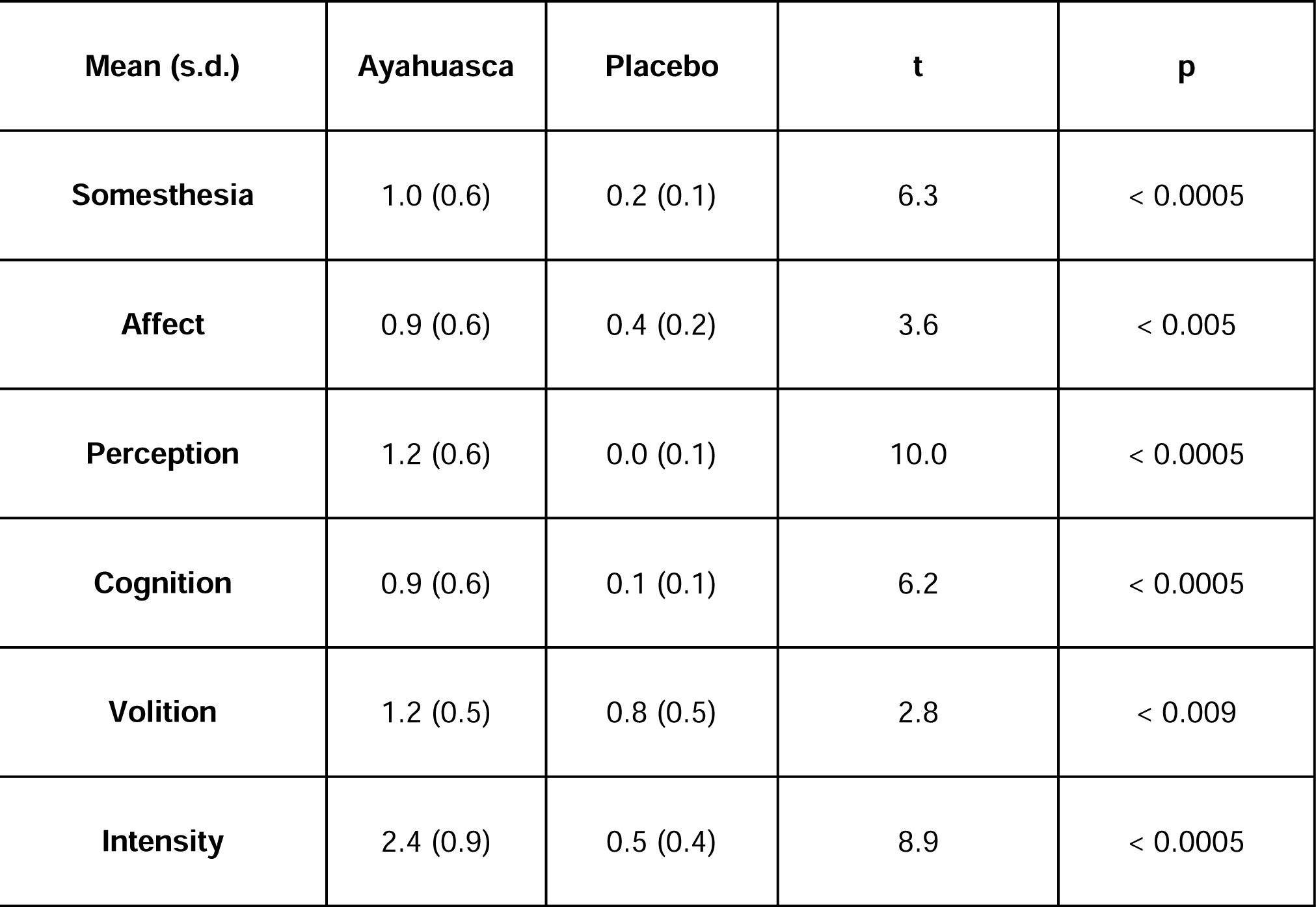
Hallucinogenic rating scale (HRS). Subscales assessed during the ayahuasca/placebo sessions. All comparisons survive multiple correction testing (p < 0.008) with the exception of the volition subscale.

### Subacute salience and default mode connectivity changes

We next explored differences in brain network functional connectivity between the ayahuasca and the placebo groups. A voxel-wise two-sample t-test revealed significant increased functional connectivity in the ayahuasca compared to the placebo group within the salience network (t = 1.3; joint cluster and extent probability thresholds of p < 0.05 and p < 0.01) (**Figure 2A** red and yellow maps, **Supplementary Table S3**). Functional connectivity increases with our seed of interest were located bilaterally within the anterior cingulate cortex and superior frontal gyrus, with a tendency towards the left hemisphere (**Figure 2A** first three brain slides and **Supplementary Table S3**). Conversely, we found functional connectivity decreases within the default mode network in the ayahuasca group compared to placebo, predominantly affecting our seed of choice in the posterior cingulate cortex (**Figure 2C** and **Supplementary Table S3**). Average levels of functional connectivity changes derived from the clusters identified in **Figures 2A** and **C** are schematized as box plots (**Figure 2B** and **D**). Similar results were derived by adding the difference in head mean framewise displacement as a covariate of no interest in the two-sample t-test models (**Supplementary Figure S2** and **Supplementary Table S4**), suggesting no influence of head motion in functional differences found across groups. Similar functional connectivity differences to our seeds of choice in the salience network and default mode network were also found when using nonparametric permutation-based tests (**Supplementary Figure S5** and **Supplementary Table S5**). Furthermore, consistent with our seed-based findings, ICA revealed patterns of increased anterior cingulate cortex and superior frontal gyrus functional connectivity for the salience network (t = 1.7; joint cluster and extent probability thresholds of p < 0.05, **Supplementary Figure S4 A-B** and **Supplementary Table S6**) and a trend in decreased posterior cingulate cortex functional connectivity of the default mode network in the ayahuasca group (t = 1.3; joint cluster and extent probability thresholds of p < 0.1, **Supplementary Figure S4 C-D** and **Supplementary Table S6**). Inter-network functional connectivity analyses (**Figure 3**) revealed increased functional connectivity between the salience and default mode networks in the ayahuasca group compared to the placebo group (t = 2.5; p < 0.02). Intra-network functional connectivity of primary sensory networks (visual and sensorimotor) and inter-network functional connectivity of primary sensory networks with the default and salience networks did not differ significantly between ayahuasca and placebo, suggesting some level of specificity of the subacute functional changes elicited by ayahuasca on the salience and default mode networks.

**Figure 2.**
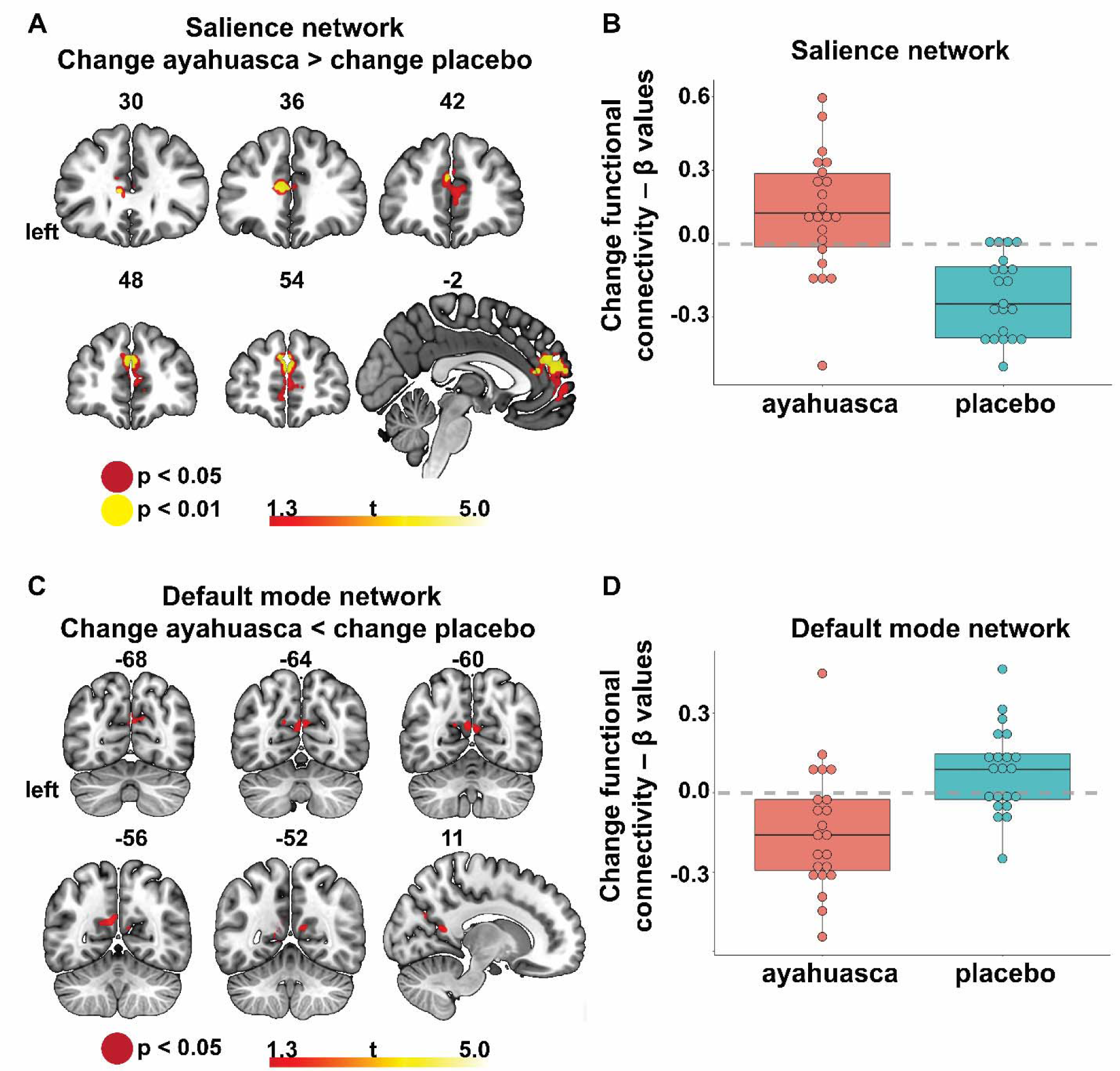
Intra-network functional connectivity changes. **(A)** Significant salience network functional connectivity increases within the anterior cingulate cortex for the ayahuasca compared to the placebo group. Joint extent and cluster probability thresholds of p < 0.05 (red) and p < 0.01 (yellow). Left is on the left, bar reflects t-values. **(B)** Box plot schematizing group differences in functional connectivity (average functional connectivity levels derived from the cluster identified in panel A with a joint cluster and extent probability thresholds of p < 0.01). **(C)** Significant default mode network functional connectivity decreases within the posterior cingulate cortex for the ayahuasca compared to the placebo group. Joint cluster and extent probability thresholds of p<0.05 (red). Left is on the left, bar reflects t-values. **(D)** Box plot schematizing group differences in functional connectivity (average functional connectivity levels derived from the cluster identified in panel C with a joint cluster and extent probability thresholds of p < 0.05).

**Figure 3.**
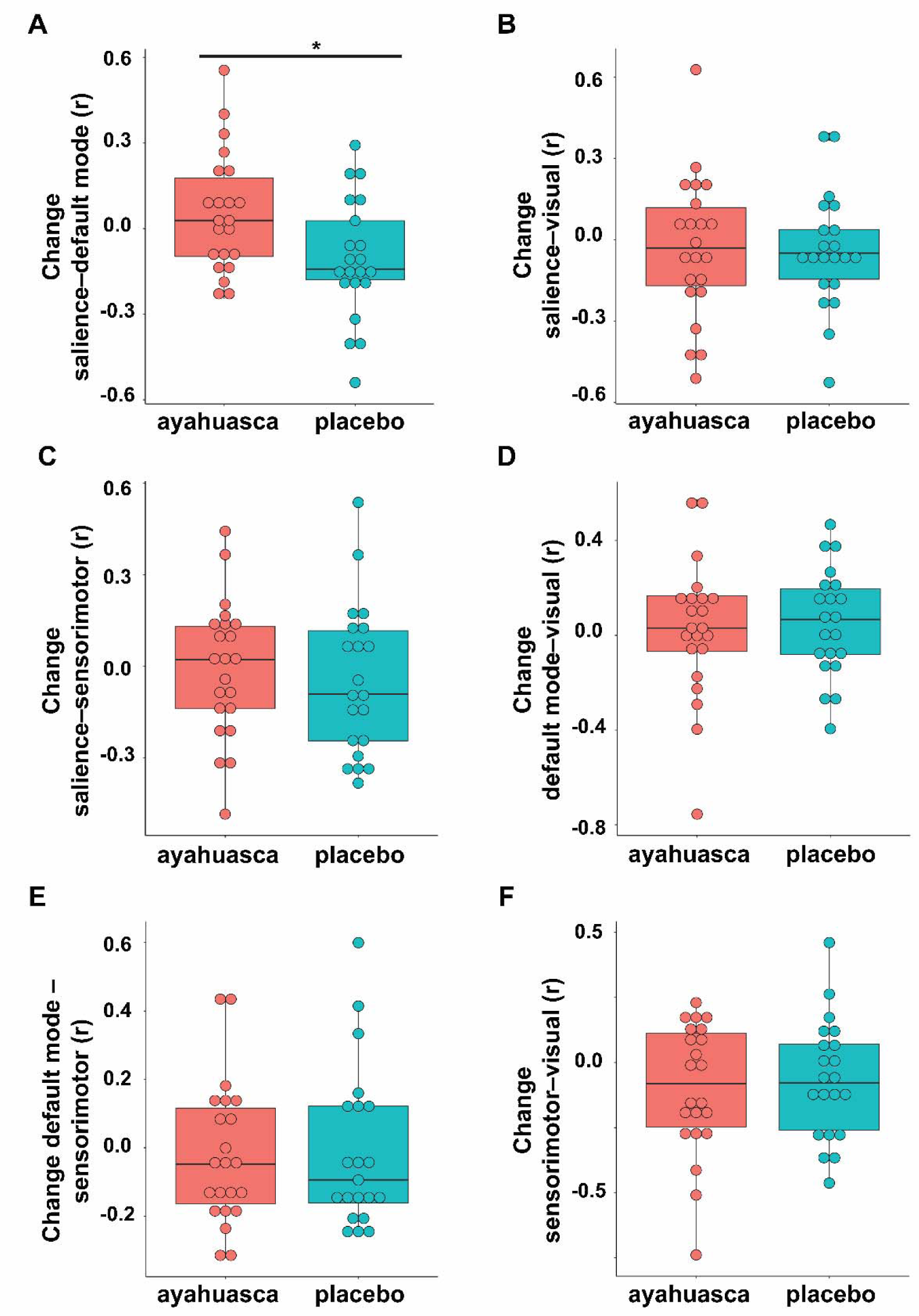
Inter-network functional connectivity changes. **(A)** Significant connectivity increases between the salience and default mode networks in the ayahuasca (in red) compared to the placebo group (in cyan). Connectivity between the **(B)** salience network and visual network, **(C)** salience network ad sensorimotor network, **(D)** default mode network and visual network, **(E)** default mode network ad sensorimotor network, **(F)** and visual network and sensorimotor network did not significantly differ between groups. r = Pearson’s correlation coefficient used to assess inter-network functional connectivity. *p < 0.02

### Ayahuasca induced subacute changes in functional connectivity correlate with acute alterations in somesthesia, affect, and volition

Average functional connectivity changes were derived from the previously identified clusters in the salience network and default mode network, and associated with HRS subscores assessed during the acute effects in an explorative way. Partial correlation analyses corrected for the difference in head mean framewise displacement between the second and first scanning session revealed significant positive associations between increases in salience network functional connectivity and somesthesia in the ayahuasca group but not in the placebo group (PR_aya_ = 0.55, p < 0.01; PR_pla_ = 0.01, p = 0.95) (**Figure 4**). Positive trend correlations were found between increased salience network functional connectivity and affect in the ayahuasca group and volition in the placebo group (**Supplementary Table S7**). Subacute changes in default mode network functional connectivity showed a significant negative correlation only with volition in the ayahuasca group and a trend in placebo (PR_aya_= - 0.59, p < 0.005; R_pla_= −0.43, p = 0.06) (**Figure 4, Supplementary Table S7**). Further, subacute changes in functional connectivity between the salience and default mode networks correlated significantly with affect in the ayahuasca but not in the placebo group (PRaya= 0.47, p < 0.03; Rpla= −0.06, p = 0.81) (**Figure 4, Supplementary Table S7**). Trend correlation were found also to cognition in the ayahuasca and to perception in the placebo group (**Supplementary Table S7**). We subsequently estimated multiple linear regression models over all participants, using somesthesia as the dependent variable, and within salience network connectivity change, the difference in head mean framewise displacement between sessions, and a categorical group variable as predictors. This analysis revealed a main effect of group (β = −0.51, t = −3.00, p < 0.005) indicating that the two slopes associating within salience network connectivity change and somesthesia scores significantly differ across the ayahuasca and the placebo groups. An analogous analysis exploring the association between affect and functional connectivity between the salience and default mode networks revealed a similar main effect of group (β = - 0.39, t = −2.64, p < 0.02) indicating significant group differences in the slopes. Using volition as the dependent variable and within default mode network functional connectivity change as predictor revealed that the main effect of group was not significant (β = −0.13, t = −0.88, p = 0.383), indicating that the slopes do not differ across groups.

**Figure 4.**
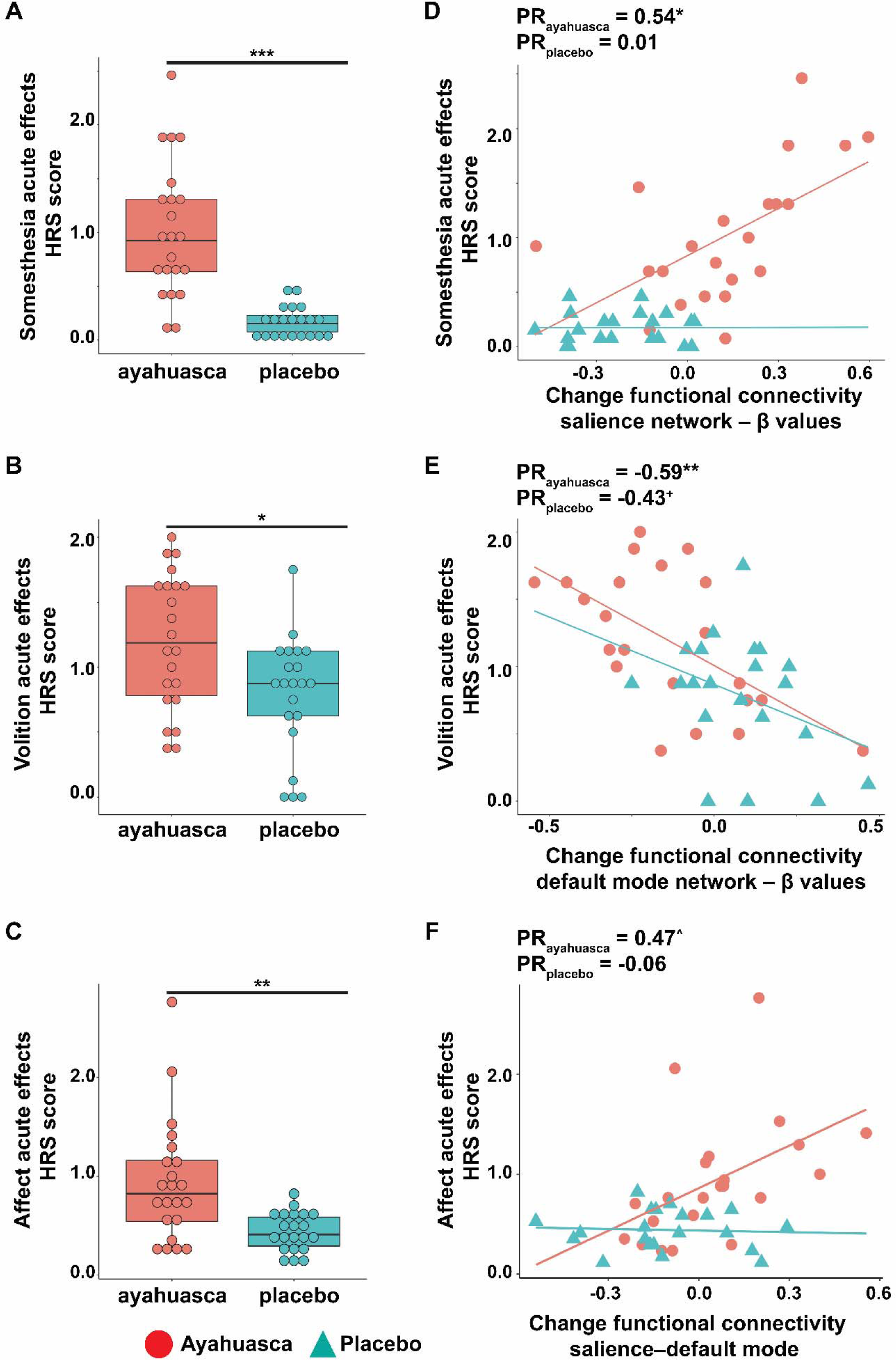
Ayahuasca induced subacute changes in functional connectivity correlate with altered somesthesia, volition, and affect. Boxplot showing group differences in **(A)** somesthesia, **(B)** volition and **(C)** affect assessed with the HRS during the acute effects of ayahuasca (in red) and placebo (in cyan). Scatter plots and partial correlation coefficients corrected for the difference in head mean framewise displacement between the second and first scan associating **(D)** somesthesia, **(E)** volition, or **(F)** affect with averaged levels of functional connectivity changes identified within the salience network, within the default mode network, and between the salience and default mode networks (ayahuasca red circles and line, placebo cyan triangles and line). _+_p < 0.1, ^p < 0.03, *p < 0.01, **p < 0.005, ***p < 0.0005.

## Discussion

Neuroimaging techniques provide a unique opportunity to study the neural correlates of altered states of consciousness induced by psychedelic agents (Carhart-Harris et al., 2012; Carhart-Harris, Muthukumaraswamy, et al., 2016; De Araujo et al., 2012; Palhano-Fontes et al., 2015; Riba et al., 2004; Viol et al., 2017; Vollenweider et al., 1997). In this study, ayahuasca naïve participants were randomly assigned to a single session with either ayahuasca or placebo, and functional connectivity of different large-scale brain networks was assessed with tf-fMRI one day before and one day after the dosing session. We found (i) salience network connectivity increases within the anterior cingulate cortex and between the anterior cingulate cortex and the superior frontal gyrus in ayahuasca compared to placebo; (ii) increased connectivity between the salience and default mode networks in the ayahuasca group; (iii) while default mode network connectivity was decreased within the posterior cingulate cortex in the ayahuasca group. Ayahuasca induced subacute functional connectivity increases in the salience network correlated with altered levels of somesthesia, which reflects somatic changes including interoceptive, visceral, and tactile effects induced by psychedelics (Riba, Rodrı, et al., 2001). Reduced default mode network connectivity correlated with altered levels of volition, proposed to reflect the subject’s capacity to willfully interact with his/her ‘self’ during the psychedelic experience (Riba, Rodrı, et al., 2001). Increased connectivity between the salience and default mode networks correlated with altered levels of affect reflecting emotional responses during the acute psychedelic session (Riba, Rodrı, et al., 2001). No subacute inter- and intra-network connectivity differences were detected in primary sensory brain networks, suggesting specificity of the observed changes in the salience and default mode networks. Although little is known on the long-term functional impact of psychedelics on large-scale brain networks, these findings suggest that ayahuasca has sustained effects on neural systems supporting interoceptive, affective, and self-referential processes (Brewer et al., 2013; Craig, 2009; Critchley and Harrison, 2013; Raichle, 2015; Seeley et al., 2007; Uddin, 2015).

### Effects of psychedelic substances on the salience network and default mode network

N,N-DMT, one of the major constituents of ayahuasca, has high affinity to the 5HT2A serotonergic receptor (Barker, 2018), while other constituents such as harmine, harmaline and tetrahydroharmine act as monoamine oxidase inhibitors and serotonin reuptake inhibitors partially by interacting with the serotonin transporter (5-HTT) (Mckenna et al., 1984; Riba, Rodríguez-Fornells, et al., 2001). A recent molecular neuroimaging study involving positron emission tomography acquired in a large sample of healthy individuals revealed *in vivo* multimodal density maps of serotonin transporter and major serotonin receptors in the human brain (Beliveau et al., 2017). 5HT2A receptors displayed a widespread pattern of distribution spanning the visual cortex, temporal, parietal and frontal regions overlapping with major hubs of the salience, default mode, and task-control networks (Buckner et al., 2008; Raichle et al., 2001; Seeley et al., 2007), with density of serotonin transporter being highest within frontotemporal regions overlapping with the insula and anterior cingulate cortex. Previous studies investigating psychedelics have reported decreased within salience network functional coupling during the acute effects of psilocybin (Lebedev et al., 2015). Psilocybin-induced reduced functional connectivity was associated with “ego-dissolution”, a construct frequently reported as altered by psychedelics, characterized by the feeling that the border between one’s self and the external world is dissolving (Goodman, 2002; Griffiths et al., 2011; Lyvers and Meester, 2012; Trichter et al., 2009). Similar studies have reported increased salience network entropy levels during the acute effects of LSD (Lebedev et al., 2016). Ketamine, an N-methyl-D-aspartate receptor antagonist, is a common dissociative anesthetic, which at subanesthetic doses has rapid and sustained antidepressant effects (Ionescu et al., 2018). In both healthy participants and patients with depression, acute ketamine administration has been shown to dampen connectivity of key regions of the salience and default mode networks such as the anterior cingulate and posterior cingulate cortices (Bonhomme et al., 2016; Lehmann et al., 2016; Scheidegger et al., 2012), whereas connectivity between these regions has been shown to increase post-session in patients (Abdallah et al., 2017). A study investigating the structural correlates of long-term ayahuasca use found increased cortical thickness within the anterior cingulate cortex (a region standing out throughout our analyses) of regular ayahuasca users compared to controls, while cortical thickness of default mode network regions such as the precuneus and posterior cingulate cortex was decreased in long-term ayahuasca users (Bouso et al., 2015). The salience network has been proposed to orchestrate dynamic switching between mentalizing states anchored on the default mode network and externally-driven attentional states anchored on the task-control network in order to guide behavior by segregating relevant internal and extrapersonal stimuli (Menon and Uddin, 2010; Uddin, 2015; Zhou and Seeley, 2014). Intriguingly, increased global coupling and functional connectivity between salience and default mode network nodes have been found under acute psilocybin administration (Carhart-Harris et al., 2013) and subacutely with ayahuasca (Sampedro et al., 2017), but future work, ideally combining multimodal molecular and functional neuroimaging, is needed in order to elucidate how mid- and long-term functional interactions of both networks are changed by psychedelic agents.

### Impact of psychedelics on interoception, affect, and self-referential processes

Psychedelic substances modulate blood pressure, body temperature, and heart rate (Holze et al., 2019) by interacting with the sympathetic and parasympathetic autonomic nervous system, and have shown to impact emotional and affective functions by improving clinical symptoms in mood disorders (Carhart-Harris, Bolstridge, et al., 2016; Griffiths et al., 2016; Grob et al., 2011; Palhano-Fontes et al., 2019; Ross et al., 2016; Sanches et al., 2016). By which mechanisms may ayahuasca mediate changes in interoception, affect, and self-referential processes? Hubs of the salience network, such as the insula and the anterior cingulate, have been consistently associated with emotional processing (Craig, 2009; Critchley and Harrison, 2013; Etkin et al., 2011; Ochsner and Gross, 2005; Seeley et al., 2007; Touroutoglou et al., 2012) and dysfunctions in these regions underlie depression and anxiety in various affective disorders (Williams, 2016). Increased connectivity in the salience network has been associated with early heightened emotional contagion in preclinical Alzheimer’s disease (Fredericks et al., 2018; Sturm et al., 2013), while widespread salience network functional and structural degeneration is observed in behavioral variant frontotemporal dementia, a neurodegenerative disease characterized by loss of empathy, socioemotional symptoms, and autonomic dysfunctions (Rankin et al., 2006; Seeley et al., 2012; Sturm, Sible, et al., 2018). Based on these studies, the salience network has been proposed to support socioemotional-autonomic processing through its interoceptive afferents in the anterior insula processing autonomic activity streams regarding the “moment-to-moment” condition of the body (Craig, 2009; Critchley and Harrison, 2013; Uddin, 2015; Zhou and Seeley, 2014). The anterior cingulate cortex subsequently receives integrated anterior insula input and serves to mobilize visceromotor responses to salient socioemotional stimuli in order to guide behavior (Craig, 2009; Critchley and Harrison, 2013; Uddin, 2015; Zhou and Seeley, 2014). Critically, our findings of subacute default mode network functional connectivity decreases with ayahuasca are in contrast with a previous study reporting subacute default mode network connectivity increases with psilocybin in treatment-resistant depression (Carhart-Harris et al., 2017). Previous works have reported both default mode network connectivity increases and decreases in major depression (Berman et al., 2011; Greicius et al., 2007; Sheline et al., 2009; Yan et al., 2019), with heightened connectivity being associated with altered self-referential thoughts such as rumination and negative internal representations (Berman et al., 2011; Sheline et al., 2009). Although in healthy subjects, our findings provide preliminary evidence that ayahuasca may have a sustained effect on dampening functional connectivity of systems involved in self-referential processes associated with the characteristic symptoms of depression (Brewer et al., 2013; Sheline et al., 2009). We propose that future studies combining longitudinal tf-fMRI and autonomic recordings in healthy and neuropsychiatric populations could shed light on the acute and long-term impact of psychedelic agents on brain networks supporting interoceptive, affective, and self-referential processes.

### Limitations

An important limitation of our study consists in the use of liberal voxel-wise statistical thresholds when assessing group differences in subacute functional connectivity across distinct brain networks. Recent research has shown that liberal cluster-extend thresholds are prone to inflate false-positive results, questioning the validity of weakly significant neuroimaging findings (Eklund et al., 2016). The low power attained in our study may have been caused by the small number of participants involved and by the moderate strength of the MRI scanner (1.5 Tesla), urging for future studies involving larger sample sizes and state-of-the-art neuroimaging acquisition. Further, weak effects on functional connectivity were to be expected after a single ayahuasca session in a sample of cognitively healthy participants. Future studies implementing longitudinal sessions and the inclusion of neuropsychiatric populations could help overcome this limitation. We performed, however, several control analyses in order to mitigate methodological and statistical concerns. First, we assessed the reliability of our findings by controlling for the used methodological approach, which revealed similar functional connectivity changes when comparing findings from the seed-based versus ICA approach. In order to address concerns related to the liberal statistical threshold, we applied nonparametric permutation tests replicating the main findings in this study. Second, we assessed the impact of head movement on intra-network functional connectivity differences across groups and on the association between altered states of consciousness and subacute functional connectivity changes through the use of partial correlation analyses. These analyses revealed that head movement in the scanner, a common confounder in tf-fMRI studies, did not significantly impact our findings. However, findings of decreased functional connectivity, particularly within the default mode network need to be interpreted with a grain of salt, since these did not survive more stringent statistical thresholds. Further, the volition subscale of the HRS has been shown to have a low internal consistency (Riba, Rodri, et al., 2001). Given the exploratory nature of the correlational analyses and the associated danger of false-positive inflation, we advise caution in interpreting the association between altered acute levels in the volition scale and subacute default mode network functional connectivity decreases induced by ayahuasca. In particular, both the ayahuasca and the placebo groups exhibited a comparable relationship between changes in default mode network functional connectivity and volition. Although studies suggest that the anterior cingulate cortex, rather than posterior brain regions, has a critical role in modulating generalized effects (Sikora et al., 2016; Wager et al., 2004), our findings could potentially reflect general placebo effects.

## Conclusions

Psychedelic substances have sustained effects on affect and self-referential processes in healthy and clinical populations days to weeks after dosing. The novelty of our study resides in elucidating the subacute effects of the psychedelic ayahuasca on functional organization of the salience and default mode networks, two brain systems distinctly involved in interoceptive, affective, and self-referential functions. While primary sensory networks did not show subacute changes in functional connectivity, increased functional connectivity of the salience network one day after the session with ayahuasca related to altered acute somesthesia levels, decreases in default mode network functional connectivity related to altered levels of volition, while salience network-default mode network connectivity increases related to altered affect levels. Our findings suggest that ayahuasca may have long-lasting effects on mood by modulating those neural circuits supporting interoceptive, affective, and self-referential functions.

## Data Availability

Data is available upon reasonable request to the corresponding author.

## Acknowledgments

L.P. was supported by the German Academic Foundation. The study was funded by the Brazilian federal agencies CNPq (grants #466760/2014 & #479466/2013) and CAPES (grants #1677/2012 & #1577/2013).

